# Application of 23 novel serological markers for identifying recent exposure to *Plasmodium vivax* parasites in an endemic population of western Thailand

**DOI:** 10.1101/2021.03.01.21252492

**Authors:** Sadudee Chotirat, Narimane Nekkab, Chalermpon Kumpitak, Jenni Hietanen, Michael T White, Kirakorn Kiattibutr, Patiwat Sa-angchai, Jessica Brewster, Eizo Takashima, Takafumi Tsuboi, Matthias Harbers, Chetan Chitnis, Julie Healer, Wai-Hong Tham, Wang Nguitragool, Ivo Mueller, Jetsumon Sattabongkot, Rhea J Longley

## Abstract

Thailand is aiming for malaria elimination by the year 2030. However, the high proportion of asymptomatic infections and the presence of the hidden hypnozoite stage of *Plasmodium vivax* are impeding these efforts. We hypothesized that a validated surveillance tool utilizing serological markers of recent exposure to *P. vivax* infection could help to identify areas of ongoing transmission. The objective of this exploratory study was to assess the ability of *P. vivax* serological exposure markers to detect residual transmission ‘hot-spots’ in Western Thailand. Total IgG levels were measured against a panel of 23 candidate *P. vivax* serological exposure markers using a multiplexed bead-based assay. A total of 4255 plasma samples from a cross-sectional survey conducted in 2012 of endemic areas in the Kanchanaburi and Ratchaburi provinces were assayed. We compared IgG levels with multiple epidemiological factors that are associated with an increased risk of *P. vivax* infection in Thailand, including age, gender and spatial location, as well as *Plasmodium* infection status itself. IgG levels to all proteins were significantly higher in the presence of a *P. vivax* infection (n=144) (t test, p<0.0001). Overall seropositivity rates varied from 2.5% (PVX_097625, merozoite surface protein 8) to 16.8% (PVX_082670, merozoite surface protein 7), with 43% of individuals seropositive to at least 1 protein. Higher IgG levels were associated with older age (>18 years, p<0.05) and males (17/23 proteins, p<0.05), supporting the paradigm that men have a higher risk of infection than females in this setting. We used a Random Forests algorithm to predict which individuals had exposure to *P. vivax* parasites in the last 9-months, based on their IgG antibody levels to a panel of 8 previously validated *P. vivax* proteins. Spatial clustering was observed at the village and regional level, with a moderate correlation between PCR prevalence and sero-prevalence as predicted by the algorithm. Our data provides proof-of-concept for application of such surrogate markers as evidence of recent exposure in low transmission areas. These data can be used to better identify geographical areas with asymptomatic infection burdens that can be targeted in elimination campaigns.

## 1 Introduction

Thailand is a region of low malaria transmission, with 5432 reported confirmed clinical cases in 2019 (data obtained from the Bureau of Vector Borne Diseases Control, Ministry of Public Health, Thailand). As malaria transmission in Thailand has declined, a change in infection dynamics has occurred with a higher proportion of infections now due to *Plasmodium vivax*. Another notable feature is that a very high proportion of infections are asymptomatic (Nguitragool et al., 2019), suggesting the number of cases reported by the World Health Organisation is likely an underestimate. This mirrors changes that have occurred in other low-moderate transmission countries, such as the Solomon Islands (Quah et al., 2019), Myanmar (Liu et al., 2019) and Laos (Lover et al., 2018), where *P. vivax* is now the dominant species and a high proportion of infections are asymptomatic.

To achieve malaria elimination, national malaria control programs now need to switch their primary focus from case management to interruption of transmission. This requires monitoring and surveillance activities to detect infections, rather than just morbidity and mortality. This includes identifying residual transmission foci that can be specifically targeted for elimination. Current surveillance tools, such as microscopy and rapid diagnostic tests, are not well suited for identifying low-density, asymptomatic infections. An additional major challenge for *P. vivax* elimination is the presence of an arrested liver-stage, known as hypnozoites, which can reactive weeks to months after the primary infection to cause relapsing infections. Relapsing infections can be responsible for over 80% of all blood-stage infections (Robinson et al., 2015;Taylor et al., 2019), thus a major gap in our diagnostic/surveillance toolkit for malaria is the inability to identify individuals that harbor hypnozoites in the absence of concurrent blood-stage infections.

Serology has been proposed as a useful means of surveillance for *P. vivax*, as antibodies are possible to measure in point-of-contact tests and can reflect recent exposure not just current infections. Multiple studies have reported on the use of serology for identifying long-term changes in *Plasmodium* transmission patterns, but the application of serology as an intervention requires more actionable information. In order to address these limitations of current surveillance tools we have recently identified and validated a panel of novel serological exposure markers for classifying individuals as recently exposed to *P. vivax* (Longley et al., 2020). We demonstrated that 57/60 tested *P. vivax* proteins were able to classify individuals as recently exposed within the past 9 months, with various levels of accuracy. Using a combination of IgG levels to 8 specific *P. vivax* proteins resulted in the highest level of accuracy (sensitivity and specificity of ∼80%), better than any single protein alone. Due to the biology of the *P. vivax* parasite, essentially all individuals harbouring hypnozoites will have had a blood-stage infection within the past 9-months (White, 2011;Battle et al., 2014). Therefore, our serological exposure markers can indirectly identify individuals with *P. vivax* hypnozoites; we demonstrated this by showing that individuals who were classified as seropositive by our algorithm went on to have recurrent *P. vivax* infections at much higher rates than those who were sero-negative (Longley et al., 2020). Our serological exposure markers that can identify recent exposure within the past 9-months with ∼80% sensitivity and specificity thus provide a novel and potentially highly informative method for identifying regions of residual transmission and individuals at the highest risk of recurrent infections.

The objective of our current study was to assess the applicability of a panel of 23 candidate *P. vivax* serological exposure markers for detecting areas of residual transmission and individuals at greater risk of infection within an endemic-area of western Thailand. We aimed to do this by measuring the association between IgG levels to the 23 *P. vivax* proteins with various epidemiological factors associated with increased risk of *P. vivax* infection, and by applying the algorithm we previously developed using IgG levels to 8 *P. vivax* proteins for predicting recent exposure of the individuals in the past 9-months.

## 2 Materials and Methods

### 2.1 Study population

We used plasma samples collected from a cross-sectional survey conducted in September 2012 at three sites of the Kanchanburi and Ratchaburi provinces in western Thailand (Bongti, Kong Mong Tha, Suan Phueng), covering a 250 km central span of the Thai Myanmar border, (Nguitragool et al., 2017). This cross-sectional survey was conducted towards the end of a period of rapidly declining malaria transmission in Thailand (Chu et al., 2020). Briefly, 4309 individuals were enrolled and a fingerprick blood sample (250 µl) was collected. Blood was separated into pellet and plasma, with both components stored at −20°C until usage. The presence of *P. falciparum* and *P. vivax* parasites was determined using 18S rRNA qPCR (Rosanas-Urgell et al., 2010;Wampfler et al., 2013). The presence of *P. vivax* infections was 3.09% and *P. falciparum* infections was 0.86% (0.26% had mixed *P. vivax/P. falciparum* infections) (Nguitragool et al., 2017). Body temperature was also recorded, and fever was defined as >37.5°C. Most *P. vivax* (91.7%) and *P. falciparum* (89.8%) infections were not accompanied by any febrile symptoms (Nguitragool et al., 2017). Demographic characteristics and behavioral risk data (such as bednet usage and travel) were obtained by trained interviewers using a structured questionnaire. *Plasmodium* infections were most common in adolescent and adult males (Nguitragool et al., 2017).

In the current study, data from 4255 individuals with epidemiological data and plasma samples available were used. Table 1 provides an overview of the demographic and epidemiological study characteristics of these volunteers.

**Table 1:** Study volunteer characteristics

| Variable | Value |
| --- | --- |
| Age (years), median (range) | 20 (0.6-92) |
| Gender, number (%) |  |
| Male | 2058 (48.37%) |
| Female | 2197 (51.63%) |
| Region, number |  |
| Bongti | 2,334 |
| Kong Mong Tha (KMT) | 407 |
| Suan Phueng | 1,514 |
| *Plasmodium* infection, number (%) | 190 (4.47%) |
| *P. vivax* | 144 |
| *P. falciparum* | 46 |
| Mixed *P. vivax/P. falciparum* | 11 |
| *P. vivax* density (copy numbers/μl), range | 0.43-11,808 |
| Fever (>37.6°C), number (%) | 28 (0.67%) |
| RDT result |  |
| Number individuals tested | 100 |
| Number individuals positive | 4 |
| Anti-malarial drugs taken in last 2 weeks, number (%) |  |
| Yes | 28 (0.66%) |
| No | 4226 (99.34%) |
| Bednet used last night/in general, number (%) |  |
| Yes | 3849 (90.5%) |
| No | 404 (9.5%) |
| Duration of bednet usage, number (%) |  |
| No bednet | 280 (6.59%) |
| 0-6 months | 449 (10.57%) |
| 6-12 months | 321 (7.53%) |
| 1-2 years | 3199 (75.29%) |
| Feeling unwell today, number (%) |  |
| Yes | 199 (4.68%) |
| No | 4059 (95.32%) |
| Taking current medications, number (%) |  |
| Yes | 294 (6.91%) |
| No | 3960 (93.09%) |

We also utilized data we had previously generated against the same *P. vivax* proteins in 274 malaria-naïve negative control samples (Longley et al., 2018a), in order to set seropositivity cut-offs for each *P. vivax* protein. These samples were obtained from 102 individuals from the Volunteer Blood Donor Registry (VBDR) in Melbourne, Australia, 72 individuals from the Thai Red Cross in Bangkok, Thailand and 100 individuals from the Australian Red Cross in Melbourne, Australia. Standard screening procedures in Thailand exclude individuals from donating to the Red Cross if they have had a confirmed malaria infection within the previous three years or have travelled to malaria endemic areas within the past year.

### 2.2 Ethics statement

Written informed consent or assent was obtained from each participant in the cross-sectional survey and, where required, their legal guardian. This study was approved by the Ethics Committee of the Faculty of Tropical Medicine, Mahidol University (Ethical Approval number MUTM 2012-044-01).

Collection and/or use of the malaria-naïve negative control samples was approved under the Human Research Ethics Committee at WEHI (#14/02).

### 2.3 P. vivax proteins

23 *P. vivax* proteins were selected based on our prior work identifying and validating candidate serological markers of recent exposure to *P. vivax* (Longley et al., 2018a). These proteins, either individually or in combination, could accurately classify individuals as infected within the last 9-months, including in individuals from a Thai cohort conducted in a similar region of western Thailand as the present study. Note that the selection was made after our first iteration of the algorithm, available as version 1 in BioRxiv (Longley et al., 2018b), with those proteins that could most accurately classify recent infection selected. We have since made further improvements to the algorithm (Longley et al., 2020). Table 2 provides details of the 23 *P. vivax* proteins used in this study. The majority of proteins (n=18) were expressed using a wheat germ cell-free (WGCF) system (CellFree Sciences, Matsuyama, Japan) at either Ehime University or CellFree Sciences. The remainder were produced using an *E. coli* expression system at the Walter and Eliza Hall Institute in Melbourne or the Institut Pasteur in Paris.

**Table 2:**
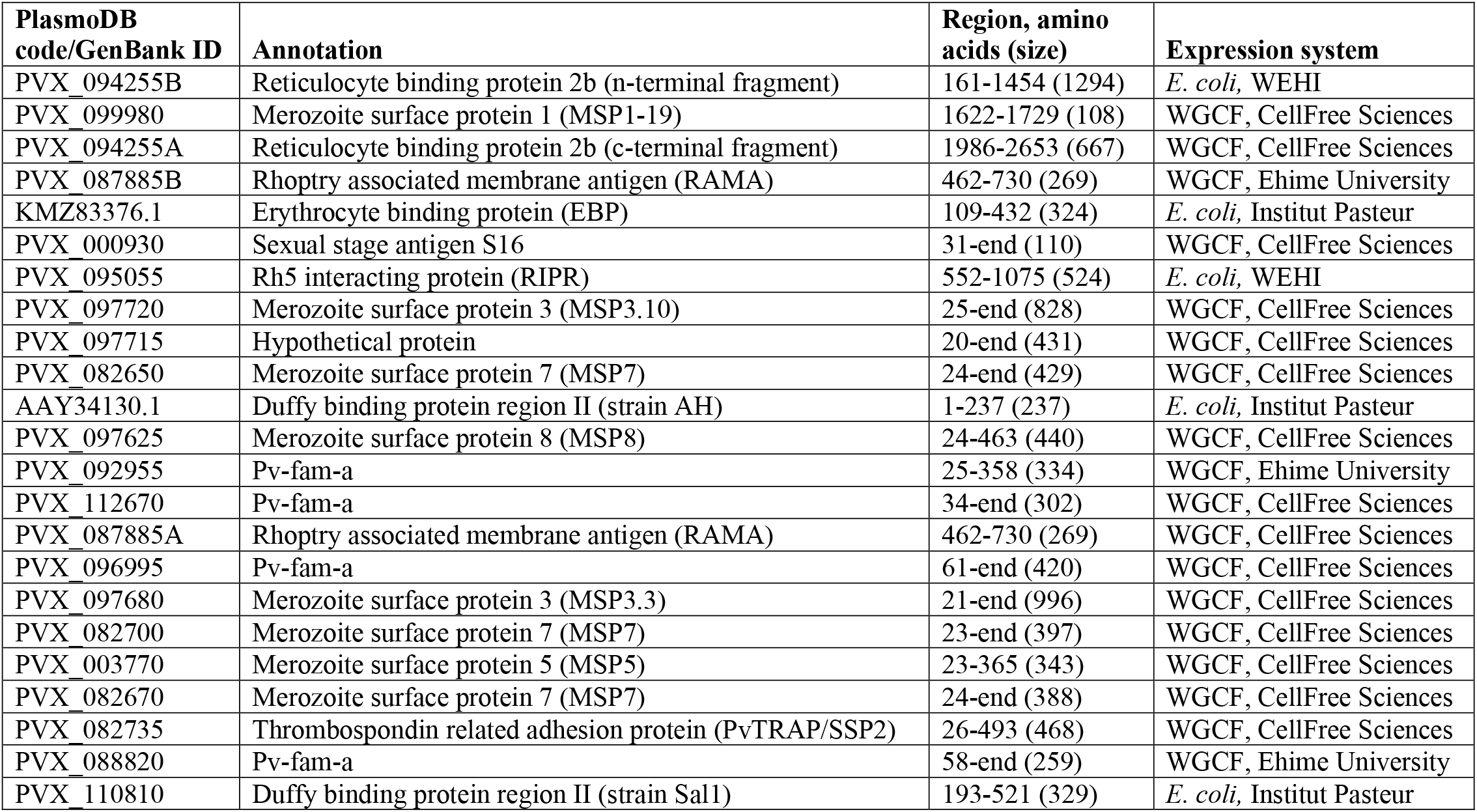
*P. vivax* proteins used in this study. A or B notation is to distinguish two constructs of the same protein and matches with our previously published protein lists (Longley et al., 2018a).

### 2.4 Multiplexed bead-based assay for total IgG measurements

Protein-specific IgG antibody levels were measured using a multiplexed bead-based assay, utilizing Luminex® technology, as previously described (Franca et al., 2016;Longley et al., 2017a). All plasma samples were run at a dilution of 1/100. Median fluorescent intensity (MFI) values from the Luminex®-200 were converted to arbitrary relative antibody units (RAU) based on a standard curve generated with a positive control plasma pool from highly immune adults from Papua New Guinea (PNG) (Franca et al., 2016). The standard curve was run on every plate. A five-parameter logistic function was used to obtain an equivalent dilution value compared to the PNG control plasma (ranging from 1.95×10^−5^ to 0.02). The conversion was performed in R (version 3.5.3).

For 21 proteins IgG levels were measured in all 4255 individuals. For proteins PVX_088820 and PVX_097715 levels were measured in 1873 and 3916 individuals, respectfully, due to availability of protein-coupled beads at the time of the study or following quality control of plate data. The complete IgG dataset generated is provided in Table S1.

### 2.5 Statistical analyses

Data analysis and presentation were performed in Prism version 8 (GraphPad, USA) or Stata version 12.1 (StataCorp, USA). Antibody values were log_10_ transformed to better fit a normal distribution. Spearman’s r correlations were used to determine the correlation between IgG levels and age. T-tests were used to determine the association between age and gender, bednet usage and the presence of *Plasmodium* infections. One-way ANOVA was used to compare IgG levels across multiple groups (i.e. regions, age categories), with adjustment for multiple comparisons performed using Sidak’s test. To determine which variables had the greatest influence on IgG levels we used a multivariable linear regression model with a stepwise backward approach, as previously described (Longley et al., 2017a). Age was included as both a linear and quadratic term, reflecting the assumption that antibody levels first increase with age but that this reaches a plateau.

A Random Forest classification algorithm was used to predict which individuals had prior exposure to *P. vivax* in the last 9 months, and were therefore likely hypnozoites carriers. The algorithm used antibody data against the top 8 *P. vivax* serological exposure markers we had previously identified (Longley et al., 2020): PVX_099980, PVX_096995, PVX_112670, PVX_087885B, PVX_097625, PVX_097720, PVX_094255B, and KMZ83376.1. The algorithm was trained with 4 datasets (Thailand (n=826), Brazil (n=925), the Solomon Islands (n=754), and negative controls (n=274)), from our previous work (Longley et al., 2020). Different diagnostic targets along the receiver operating characteristic (ROC) curve of the algorithm could be selected (Figure S1): we selected a threshold with 62% sensitivity and 90% specificity given the PCR prevalence in this study was low (thus a high specificity target was prioritised to reduce the number of potential false positives). Since the performance of our validated diagnostic tests are known, we can account for potential misclassification bias due to imperfect sensitivity and specificity in classifying individuals in our cross-sectional study. Given that the measured sero-prevalence (M) is a function of the true sero-prevalence (T) and the sensitivity (SE) and specificity (SP) of our diagnostic test, we can derive the following equation:

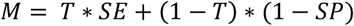

We can rearrange our equation to solve for the true (T) or, in this case, the adjusted (A) sero-prevalence, which we expect to be very close to the true sero-prevalence:

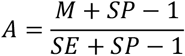

It should be noted that this does not account for relapse periodicity, lifetime exposure, immunity, and other factors that contribute to the uncertainty of these estimates.

## 3 Results

### 3.1 Immunogenicity and seropositivity of the IgG response against 23 candidate *P. vivax* serological exposure markers

We first determined population levels of immunogenicity or seropositivity against the 23 *P. vivax* proteins, using two distinct methods.

The multiplexed bead-based assay is run with a reference standard curve generated from a pool of plasma from hyperimmune individuals from PNG. We assessed the proportion of our Thai volunteers who exceeded the IgG level of the PNG pool (relative antibody units exceeding equivalent of 1/100 dilution, i.e. 0.01), to determine the proportion of individuals who have highly immunogenic responses. This varied from 0.8% (PVX_097625, merozoite surface protein 8 (MSP8)) to 9.96% (PVX_097715, hypothetical protein) across the 23 *P. vivax* proteins (Figure 1A).

**Figure 1:**
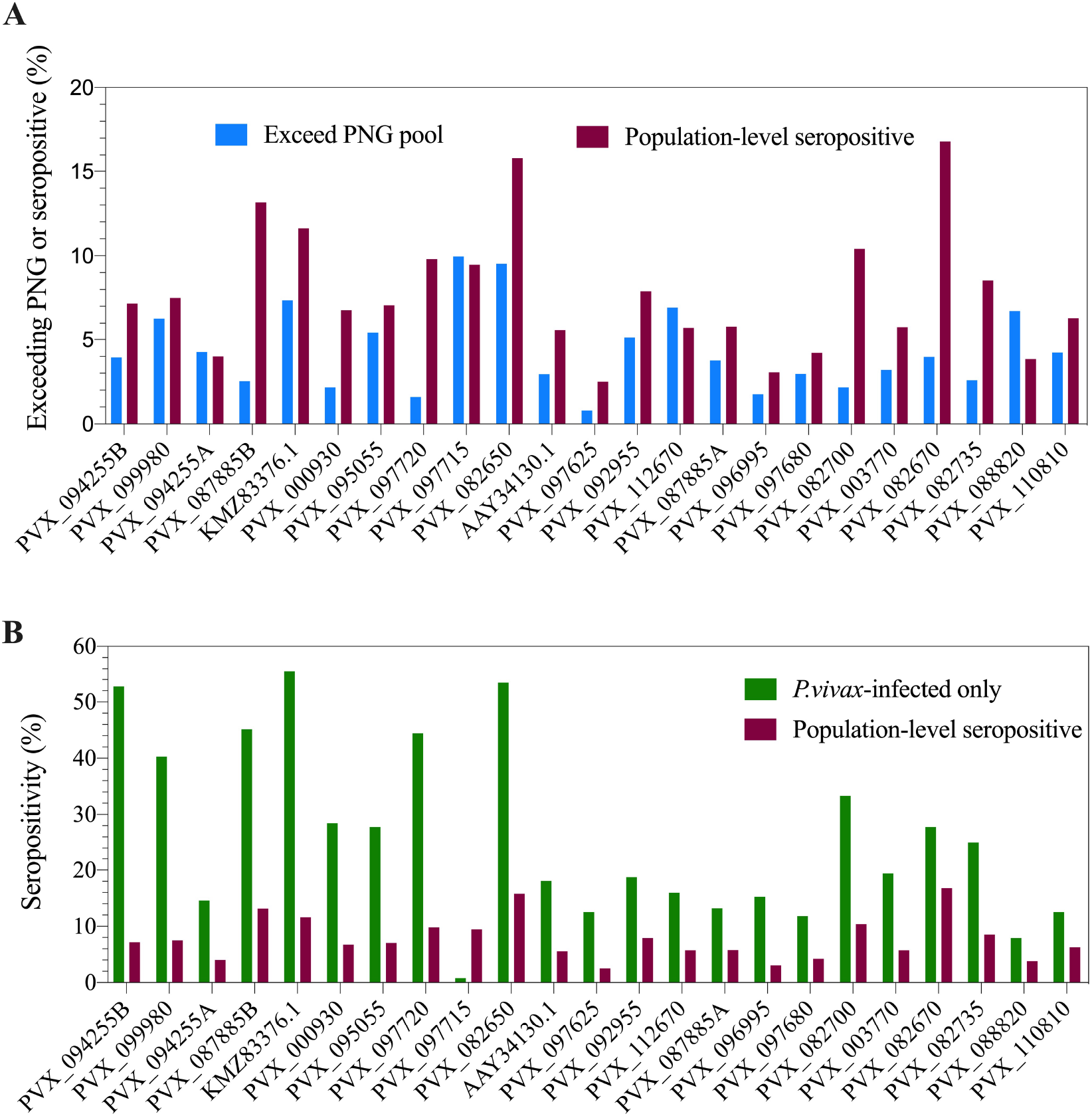
Population-level seropositivity and immunogenicity of the IgG response against 23 *P. vivax* proteins. IgG levels were measured in 1873-4255 individuals from a 2012 Thai cross-sectional survey. A) The percentage of individuals exceeding equivalent of 1/100 of the PNG pool was calculated using RAU values for each protein. The percentage of individuals seropositive was calculated by comparing to seropositivity cut-offs, defined as the mean + 2x the standard deviation of the negative control samples (n=274). B) Population-level seropositivity rates (n=1873-4255) compared to seropositivity rates in individuals with current *P. vivax* infections (n=76-144).

To determine seropositivity rates we set a protein-specific seropositivity cut-off using IgG values (relative antibody units) we previously obtained against a panel of 274 malaria-naïve negative control samples (from Melbourne, Australia and Bangkok, Thailand). The cut-off was set at the mean plus two-times the standard deviation. Seropositivity rates varied from 2.5% (PVX_097625, MSP8) to 16.8% (PVX_082670, MSP7) (Figure 1A). As the transmission level is low in this region of western Thailand (4.47% prevalence for any *Plasmodium* infection in this study), we also calculated seropositivity rates in the subset of individuals who had concurrent *P. vivax* infections during the cross-sectional survey (n=76-144, dependent on protein). This resulted in an increase in seropositivity rates for nearly all proteins, with the maximum reaching 56% of individuals seropositive against *P. vivax* erythrocyte binding protein (EBP, KMZ83376.1) (Figure 1B).

The breadth of the response per individual was also calculated, based on how many proteins each individual was seropositive to. This was calculated for 21 of the 23 *P. vivax* proteins that had complete data (PVX_097715 and PVX_088820 were excluded due to incomplete data, as detailed in the Materials and Methods). The breadth of the response per individual ranged from 0 to 21 proteins. Overall, 43% of individuals were seropositive to at least 1 protein; this increases to 90% for individuals with a concurrent *P. vivax* infection.

### 3.2 Association of IgG level with individual demographic variables known to relate to the risk of malaria infection

A number of individual-level demographic variables are known to be associated with risk of malaria infections, including age, gender and bednet usage. In particular, males were shown to have a higher risk of *Plasmodium* exposure in the individuals from the current cross-sectional survey (Nguitragool et al., 2017), and IgG levels are generally expected to increase with age in malaria-endemic regions (Longley et al., 2017a) due to increasing chance of exposure to the parasite.

We observed a weak but statistically significant correlation between IgG levels and age for all 23 *P. vivax* proteins (Spearman’s r 0.13-0.59, p<0.0001). The clearest distinction was between adults (> 18 years of age) and children (< 18 years of age) (Figure 2A), with a significant difference between these 2 groups for 19/23 proteins (One-way ANOVA with Sidak’s multiple comparison test, p<0.0001). For 18 of 23 *P. vivax* proteins there was a significantly higher IgG level in males than females (T-tests, p<0.05) (Figure 2B).

**Figure 2:**
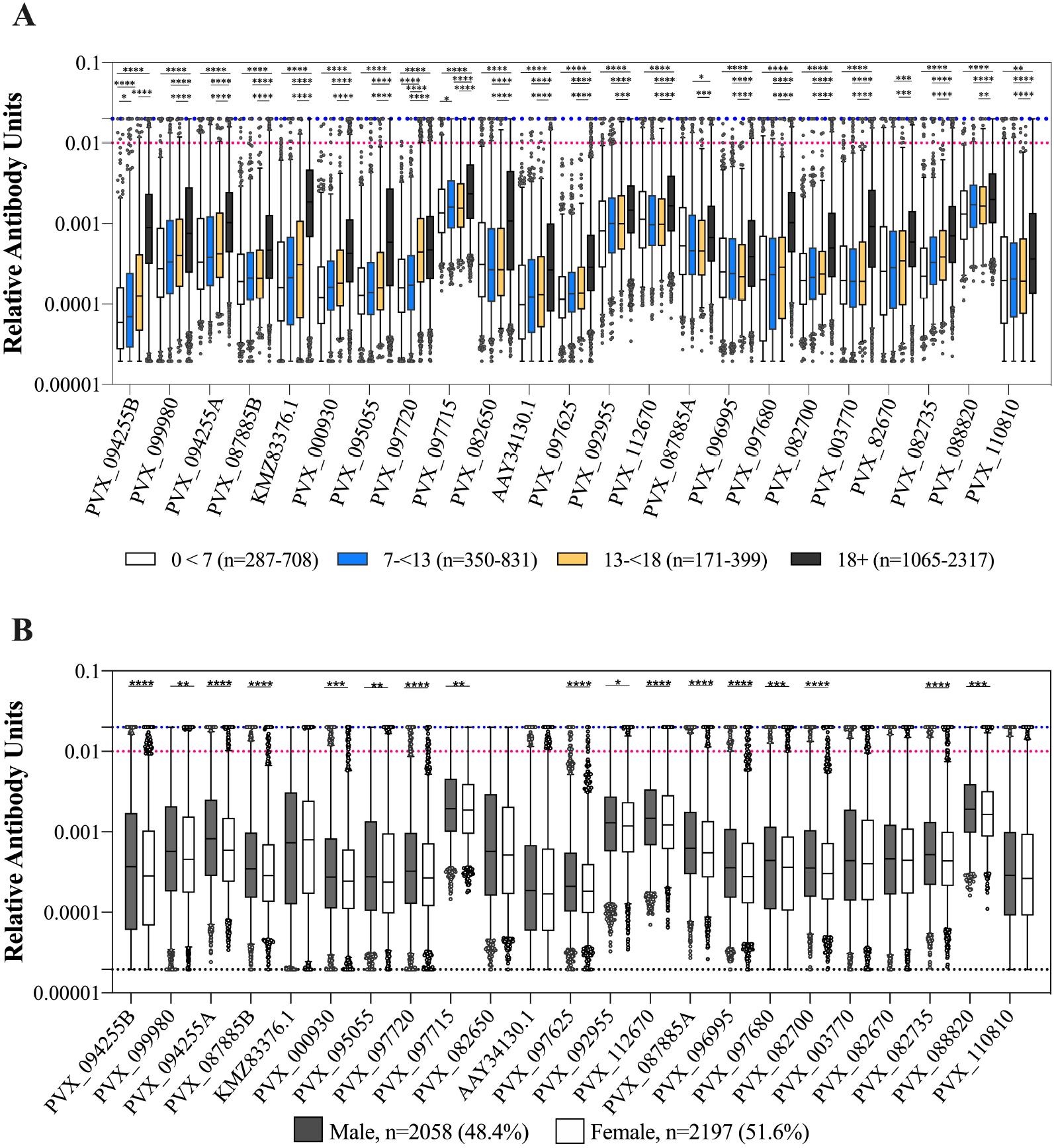
Association of IgG levels against 23 *P. vivax* proteins with age and gender. A) IgG levels compared between individuals from the 2012 cross-sectional survey as split by age: 0-7, 7-13, 13-18 and more than 18 years. Statistical significance between groups was assessed by One-way ANOVA with Sidak’s multiple comparison test. B) IgG levels compared between gender of participants (male or female). Statistical significance was assessed by T-test. The black dashed line indicates the assay minimum, the blue dashed line the assay maximum, and the red dashed line is equivalent to a 1/100 dilution of the PNG pool. * p<0.05, ** p<0.01, *** p<0.001, **** p<0.0001

We also assessed whether IgG levels varied with reported bednet usage (defined as bednet used last night/in general as yes or no). There was a significantly higher IgG level in individuals who reported using a bednet compared to those who did not for 9 of 23 *P. vivax* proteins (39%) (PVX_112670, PVX_003770, PVX_087885, PVX_097680, PVX_082670, PVX_097720, KMZ83376.1, PVX_110810, PVX_092995). The strongest association was for the proteins PVX_097680 (MSP3) and KMZ83376.1 (EBP) (T-tests, p<0.0001). There was a significantly lower IgG level in individuals who reported using a bednet for 1 protein only, PVX_088820 (a Pv-fam-a protein) (p=0.0019) (noting that this was the protein with the smallest sample size, n=1873). In the epidemiological analysis of the cross-sectional survey data, bednet usage was not associated with a reduction in risk of infection (Nguitragool et al., 2017).

### 3.3 Association of IgG levels with *P. vivax* and *P. falciparum* infection status

190 individuals (4.4% of total 4309 individuals) in the cross-sectional survey had a current *Plasmodium* infection at the time of sampling (determined by qPCR) (Nguitragool et al., 2017). The majority of infections were caused by *P. vivax* (n=144, 3.3%), with the remainder due to *P. falciparum* (n=46, 1.1%). 11 of these infected individuals had mixed *P. falciparum/P. vivax* infections. Samples were not assayed for other species of *Plasmodium*. As these were cross-sectional samples we do not know when the individuals acquired the *P. vivax* infection, but note that the average duration of an (untreated) *P. vivax* infection in Thailand has been estimated as relatively short (29 days) (White et al., 2018).

We assessed how the IgG level differed in individuals with or without *Plasmodium* infections (caused by either *P. vivax* or *P. falciparum*). As expected, there was a statistically significantly higher IgG level in individuals with a current *P. vivax* infection compared to those without for all proteins tested (p<0.0001) (Figure 3A). Interestingly, the same relationship was also evident for individuals with *P. falciparum* infections (p<0.001) (Figure 3B). Notably, 9 of the 23 *P. vivax* proteins do not have *P. falciparum* orthologs, suggesting this effect is unlikely to be due to cross-reactivity. The higher IgG levels in individuals with *P. falciparum* infection remained evident even when the 11 individuals that were co-infected with *P. vivax* were excluded (p<0.01). This finding is likely attributable to shared epidemiological risk factors for *P. vivax* and *P. falciparum* exposure.

**Figure 3:**
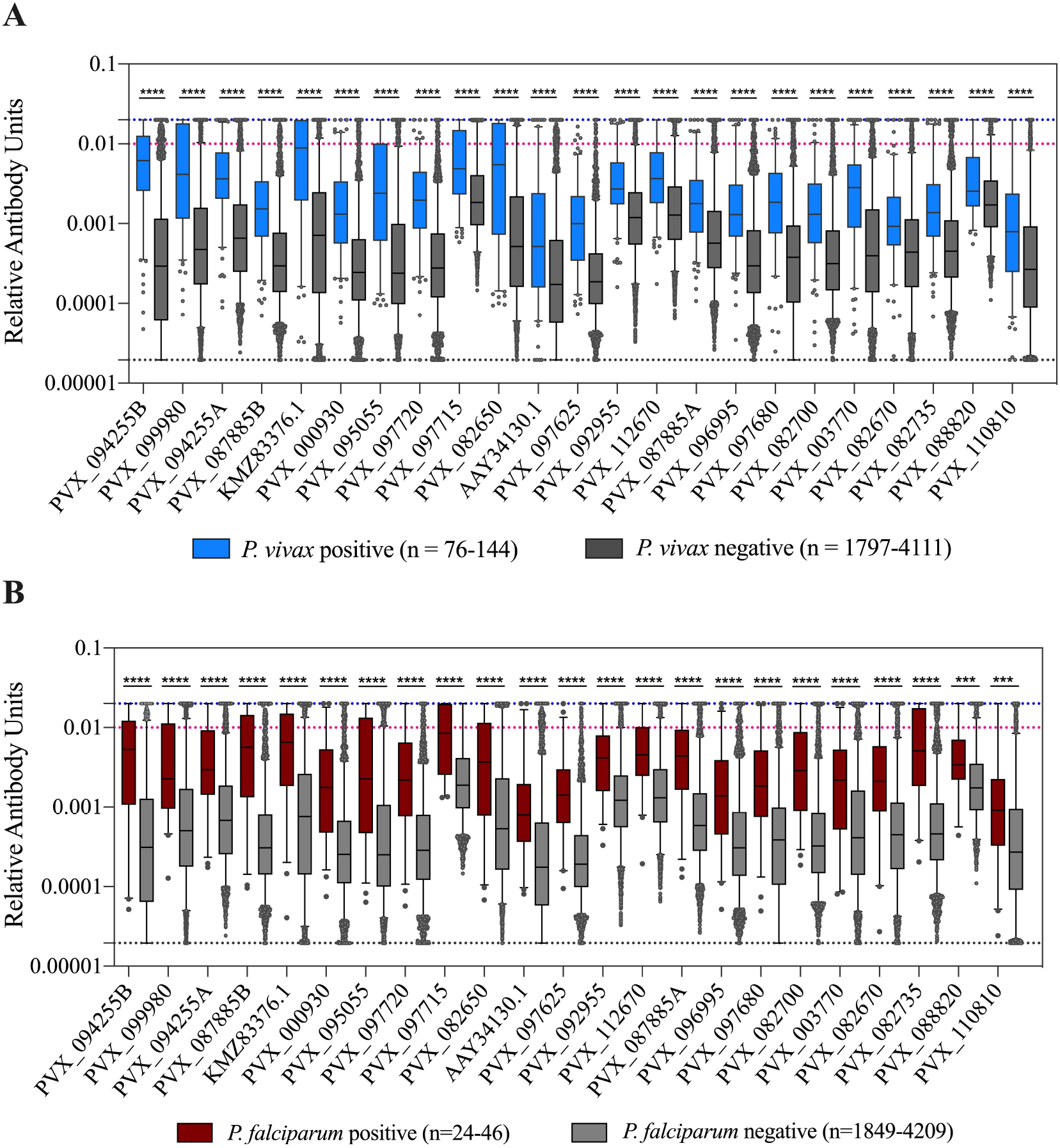
Association of IgG levels against 23 *P. vivax* proteins with concurrent *P. vivax* or *P. falciparum* infections. A) IgG levels are shown against the 23 proteins in individuals with or without current *P. vivax* infections. B) IgG levels are shown against the 23 proteins in individuals with or without current *P. falciparum* infections. Note these include individuals with *P. falciparum*-*P. vivax* co-infections. The black dashed line indicates the assay minimum, the blue dashed line the assay maximum, and the red dashed line is equivalent to a 1/100 dilution of the PNG pool. Statistical significance was assessed using T-tests. * p<0.05, ** p<0.01, *** p<0.001, **** p<0.0001

In individuals with *P. vivax* infections, we also assessed whether there was a relationship between parasite density and IgG level. There was no correlation evident between parasite density (measured as number of copies/µl blood by PCR) and IgG levels for all *P. vivax* proteins (p>0.05), suggesting that current antigenic density was not affecting the magnitude of the IgG response.

### 3.4 Association of IgG levels with spatial household location of study individuals

Individuals enrolled in this cross-sectional survey lived in 3 main regions: Bongti (3 villages), Kong Mong Tha (1 village) and Suan Phueng (4 villages). These regions have stable populations, with less than 2% of individuals reporting travel in the previous months (Nguitragool et al., 2017). They are all situated on the Thai-Myanmar border but in distinct areas over a 250 km span. There was evidence of spatial heterogeneity in terms of which individuals in the survey had *P. vivax* infections (i.e. evidence of spatial clustering or hot-spots of infection) (Figure 4), with similar *P. vivax* prevalence rates of 3.82% and 3.12% in Bongti and Suan Phueng, respectively, but much lower at 1.45% in Kong Mong Tha (Nguitragool et al., 2017). There was also variation in *P. vivax* prevalence rates amongst the villages within the 3 study sites, which is also depicted in Figure 4.

**Figure 4:**
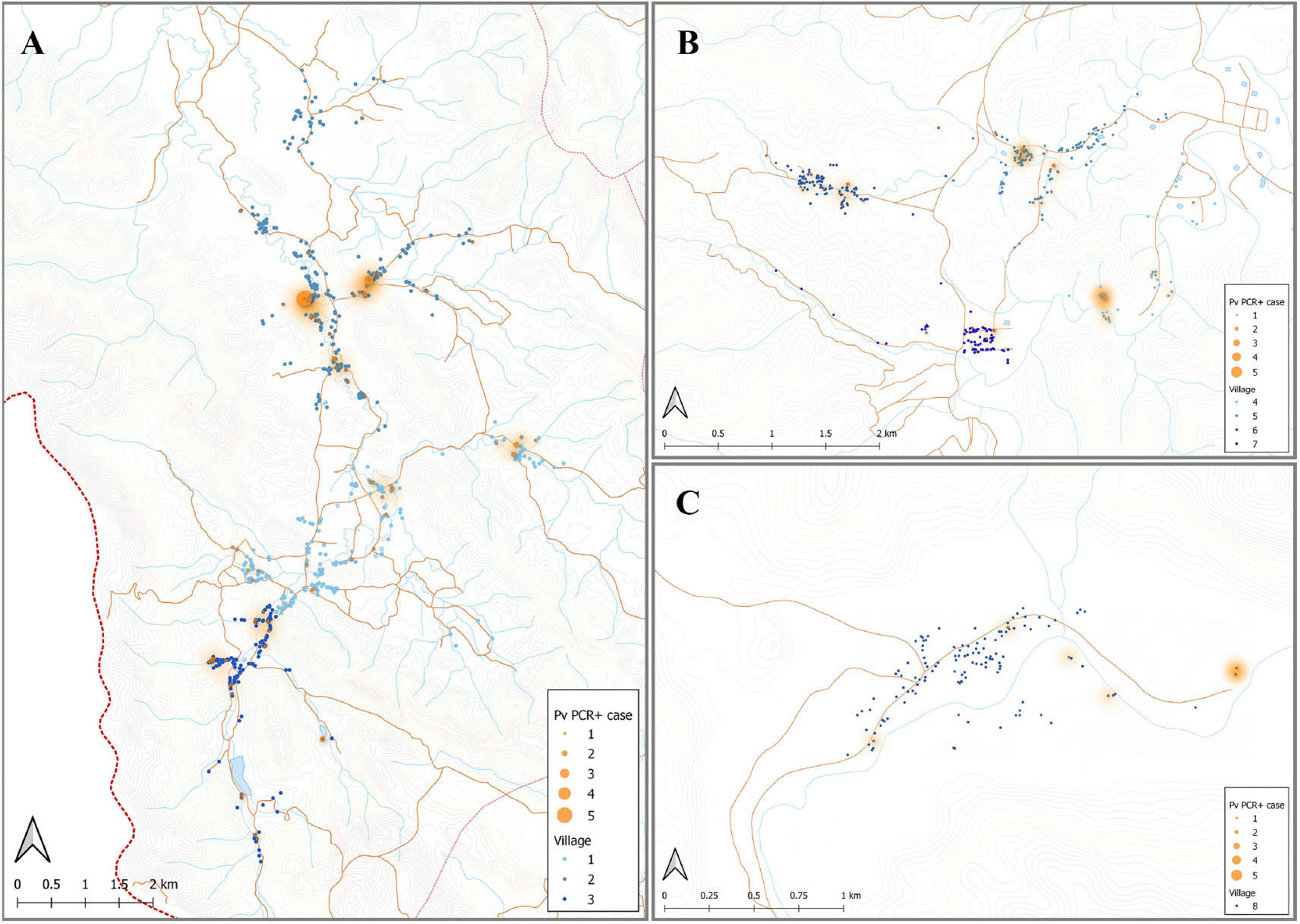
Spatial distribution of *P. vivax* infections in the 2012 cross-sectional survey. The location of individuals (at the household level) with *P. vivax* infections (orange) in 8 villages of Bongti (A), Suan Phueng (B) and Kong Mong Tha (C). The size of orange dots represents the PCR positive cases ranging from 1 to 5. The shading in orange represents multiple positive individuals in the same household or multiple positive households in the one area. Blue dots are indicative of uninfected individuals. The map was made using QGIS 3.16 software. Bongti had the largest number of enrolled individuals (n=2,334) including village 1: Ban Bong Ti Bon with 31 with *P. vivax* infections, village 2: Ban Bong Ti Lang with 37 with *P. vivax* infections, and village 3: Ban Thai Muang with 22 with *P. vivax* infections. Suan Phueng was the second largest region with 1,514 enrolled individuals, including village 4: Wangko with 14 *P. vivax* infections, village 5: Huai Krawan 14 with *P. vivax* infections, village 6: Pong Hang with 13 *P. vivax* infections, and village 7: Huai Phak with 7 *P. vivax* infections. Kong Mong Tha (village 8) was the smallest region (n = 407 enrolled individuals) and also had the lowest number of *P. vivax* infections: 6 total.

We used a one-way ANOVA to determine whether we could also identify differences in IgG levels between the three main regions. Protein PVX_088820 was excluded as all IgG data obtained was from individuals living in Bongti only. For all remaining proteins there was a statistically significant difference between groups (p<0.05) (Table 3), with the exception of PVX_094255A (RBP2b_1986-2653_). Using pairwise comparisons and adjusting for multiple testing, 5 proteins induced significantly different IgG levels in all 3 regions (PVX_096995, PVX_097680, PVX_097625, PVX_110810, PVX_095055), whilst the most common pattern was for significant differences between Bongti compared to both Kong Mong Tha and Suan Pheung (evident for 13 proteins). The trend was for the highest IgG levels in Bongti followed by Suan Pheung then Kong Mong Tha, reflecting the distribution of *P. vivax* infections. We further assessed the difference in *P. vivax-*specific antibodies between villages once we had used the IgG levels to the validated subset of 8 *P. vivax* proteins to classify individual as recently exposed to *P. vivax* or not, as this reduced the information to one variable for the antibody response rather than 21.

**Table 3:**
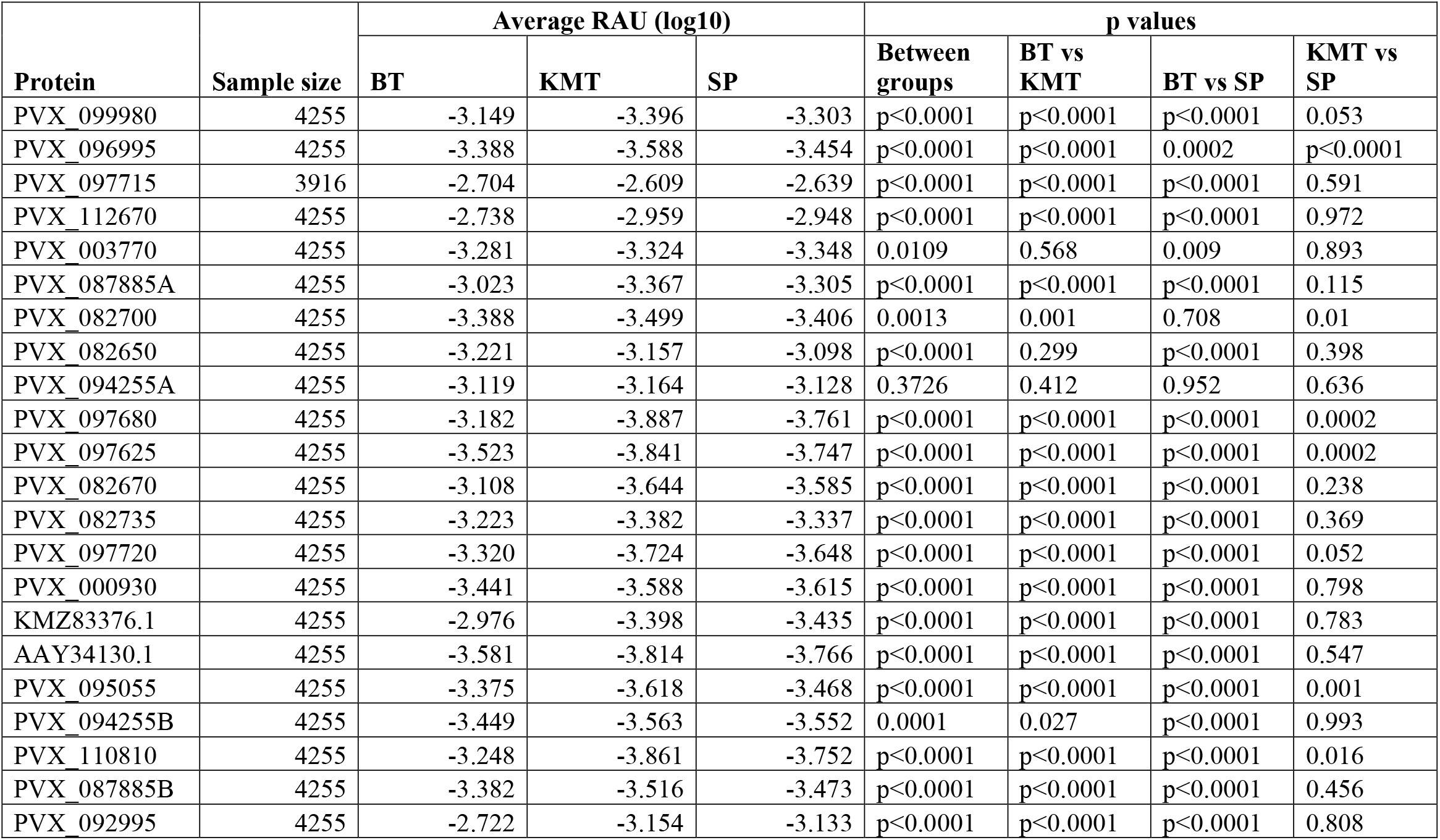
Association between IgG level to the *P. vivax* proteins with location of household. RAU = Relative Antibody Units, BT = Bongti, SP = Suan Phung, KMT = Kong Mong Tha.

### 3.5 Key variables influencing IgG level

We identified multiple variables that were associated with differences in IgG levels to the *P. vivax* proteins: age, gender, bednet usage, *Plasmodium* infections and spatial location. We therefore used a multivariable regression model to identify the variables that were having the highest influence on IgG levels. For nearly all *P. vivax* proteins, age, gender, *P. vivax* and *P. falciparum* infections and spatial location all had significant influences on IgG levels in the final multivariable model (using a backward stepwise approach as detailed in the Materials and Methods). A number of proteins had a slightly different combination of variables in the final model as detailed in Table S2. *Plasmodium* infections had the biggest impact on varying the IgG level.

### 3.6 Predicting hypnozoite carriers

We used the antibody responses generated to our previously identified top 8 *P. vivax* serological exposure markers (PVX_099980, PVX_096995, PVX_112670, PVX_087885, PVX_097625, PVX_097720, PVX_094255B, and KMZ83376.1) to predict which individuals had exposure to *P. vivax* in the previous 9 months, and could therefore potentially be harbouring hypnozoites in their livers. This also enabled us to reduce multiple antibody levels to one variable; whether or not the individuals were predicted sero-positive or sero-negative according to the algorithm. As described in the Materials and Methods, since PCR prevalence in the study was low, we also chose a high specificity target to reduce the number of potential false positives that may contribute to the over-estimation of sero-prevalence.

Using a 62% sensitivity and 90% specificity target, the algorithm identified 644 sero-positive individuals with an estimated 15.6% (95% CI 14.5%-16.7%) sero-prevalence (Table 4). This diagnostic target identified 75% of currently infected individuals, i.e. 108 out of 144 PCR+ cases. This target identified an additional 556 potential hypnozoites carriers (Table 4). This suggests a potential 17% of individuals (144 + 556) in this cross-sectional survey are either currently infected (in the blood or the liver) or have had recent past exposure to *P. vivax*. Given we know the sensitivity and specificity of our test, we can calculate an adjusted sero-prevalence using the formula in the Materials and Methods. With the high specificity target at 90% and 62% sensitivity, an adjusted sero-prevalence of 10.8% is calculated.

**Table 4:**
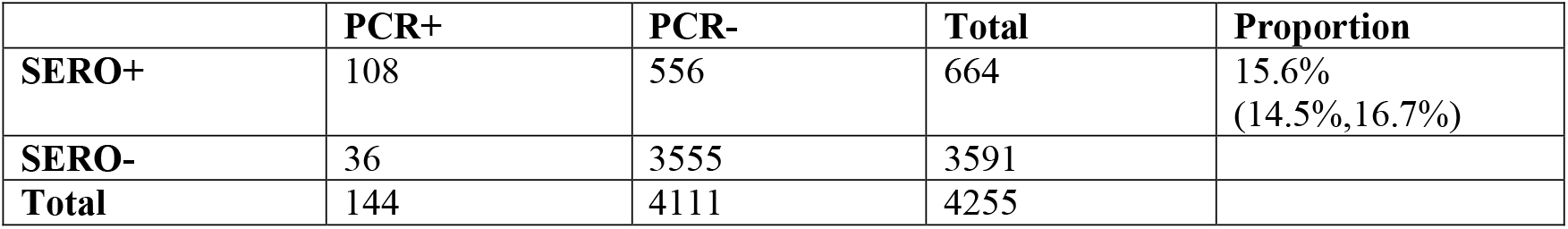
Classification with a 62% sensitivity and 90% specificity target.

Using the classification as sero-positive or sero-negative from this algorithm (target of 62% sensitivity, 90% specificity), we generated a second spatial map that demonstrates clustering of sero-positive households (Figure 5). We observed the highest levels of sero-prevalence from the algorithm in Suan Phueng (17.1%, adjusted level is 13.7%), followed by Bongti (15.1%, adjusted 10.0%) then Kong Mong Tha (12.5%, adjusted 4.9%). Kong Mong Tha also had the lowest PCR prevalence (1.47%, see Figure 4), however the highest PCR prevalence was observed in Bongti (3.86%, compared to 3.17% in Suan Phueng). At the village level, there was a significant moderate correlation between PCR prevalence and adjusted sero-prevalence (pearson r = 0.73, p = 0.038, n= 8 villages) (Figure 6).

**Figure 5:**
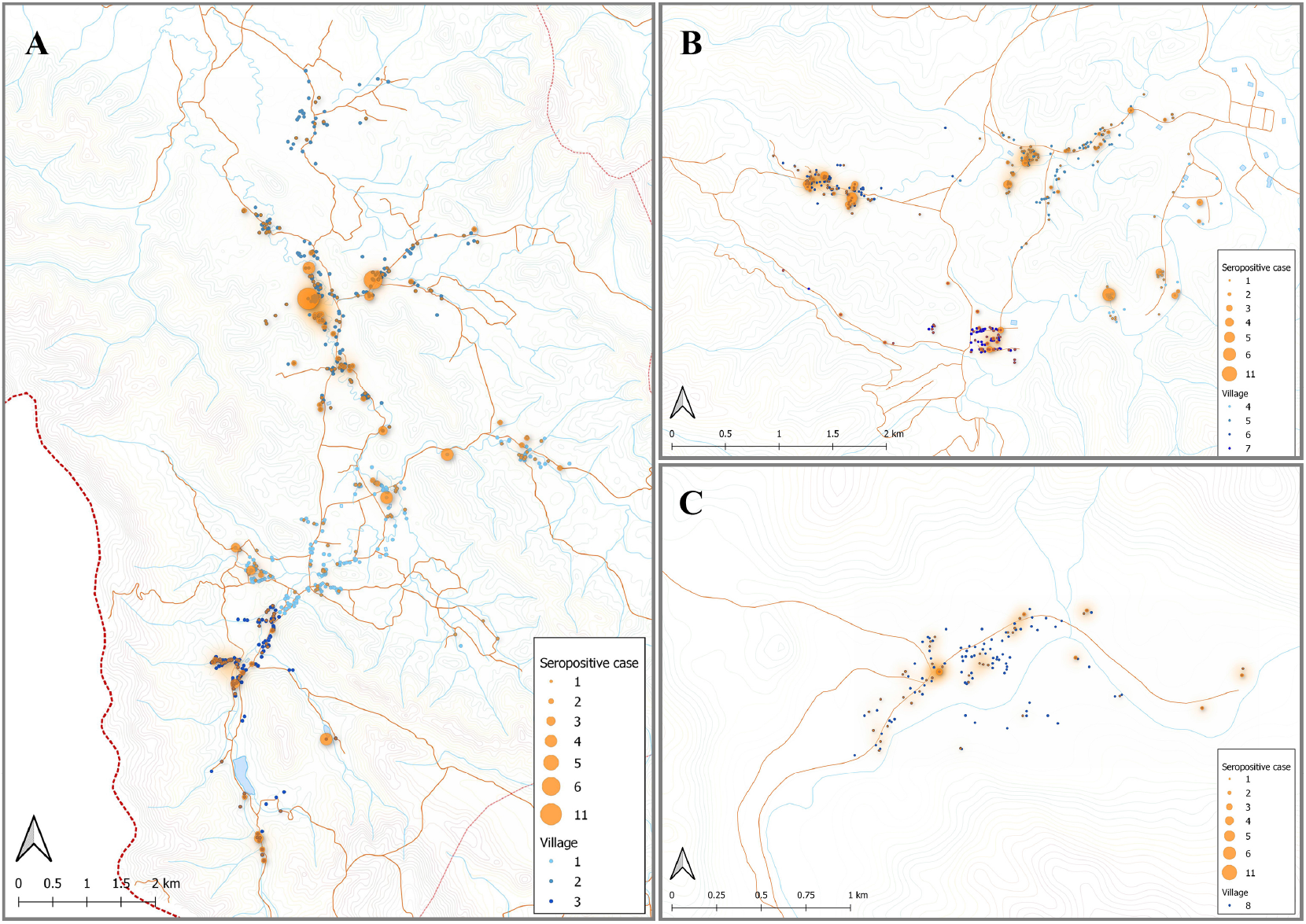
Spatial distribution of model predicted recent *P. vivax* exposure status. The location of individuals (at the household level) who were classed as seropositive (orange dot) in the algorithm, using the antibody responses against the top 8 *P. vivax* proteins. The size of orange dots represents the seropositive cases ranging from 1 to 11. The shading in orange represents multiple sero-positive individuals in the same household or multiple positive households in the one area. Blue dots are indicative of seronegative in the algorithm. The map was made using QGIS 3.16 software. Bongti (A) had 354 seropositive individuals out of a total 2334 individuals tested (sero-prevalence of 15.2%). Suan Phueng (B) was the second largest region with seropositive prevalence of 259 in the 1514 individuals (17.1%). Kong Mong Tha (C) was the smallest region and also had the lowest sero-prevalence, with 51 of the 407 individuals having a serological signature of recent *P. vivax* exposure (12.5% sero-prevalence).

**Figure 6:**
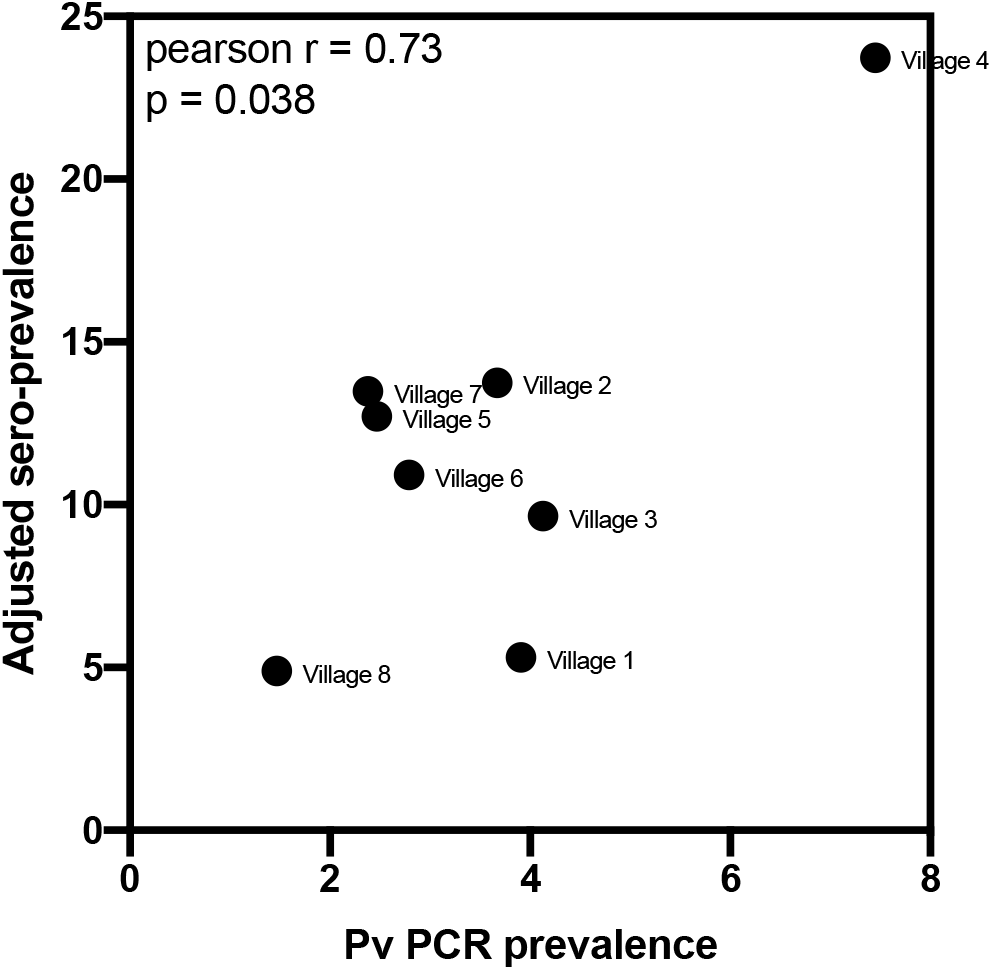
Association between village-level PCR prevalence and adjusted *P. vivax* sero-prevalence. *P. vivax* sero-prevalence was predicted by the algorithm incorporating antibody responses to the panel of 8 *P. vivax* proteins, using the 62% sensitivity, 90% specificity target. The estimates were adjusted, at the village-level, for to account for the known sensitivity and specificity. A pearson r correlation was performed to assess the association between PCR prevalence and adjusted *P*.*vivax* sero-prevalence. Data passed the Anderson-Darling test for normality.

## 4 Discussion

Serological exposure markers for identifying regions of residual or ongoing transmission of *P. vivax* could play a unique role in control and elimination efforts of malaria-endemic countries in the Asia-Pacific (and elsewhere). In this study, we measured total IgG antibody responses against a panel of 23 *P. vivax* proteins that we have previously shown to be accurate markers of recent exposure to *P. vivax* parasites. By associating total IgG responses with the presence of *Plasmodium* infections, in addition to various epidemiological risk factors of exposure, we provide further evidence that these responses can be used as markers of residual transmission.

The majority of individuals surveyed in this cross-sectional study had no detected *Plasmodium* infections by qPCR. Accordingly, the seropositivity rates against the 23 *P. vivax* proteins were low, ranging from 2.5% (PVX_097625, MSP8) to 16.8% (PVX_082670, MSP7). The seropositivity rates increased when we limited our analysis to the 144 individuals with current *P. vivax* infections, reaching a maximum of 56% against the protein KMZ83376.1 (EBP). These seropositivity rates are very similar to what we have previously observed in a longitudinal cohort study conducted in the Kanchanaburi province (Longley et al., 2018a). Of the 23 *P. vivax* proteins, our top 8 proteins as previously identified were PVX_094255 (reticulocyte binding protein 2b, RBP2b), PVX_087885 (rhoptry associated membrane antigen, RAMA), PVX_099980 (MSP1), PVX_096995 (a Pv-fam-a protein), PVX_097625 (MSP8), PVX_112670 (an unknown protein), KMZ83376.1 (EBP) and PVX_097720 (MSP3). In the entire study set of this cross-sectional survey the seropositivity rates to the top 8 were similar to the rest of the *P. vivax* proteins, however in the individuals with current *P. vivax* infections 5 of the top 8 proteins exhibited the highest levels of seropositivity (>40%). Whilst we do not know when the individuals in this cross-sectional survey acquired the *P. vivax* infection, the relatively high levels of seropositivity against our markers in individuals with current infections is a promising feature. Importantly, some individuals with current PCR-confirmed infections may have only just acquired the infection, and thus the antibody response would not be expected to peak for another 1-2 weeks. An additional protein not within our top 8 that had a high seropositivity rate in individuals with current *P. vivax* infections was MSP7, with 53% seropositivity. The selection of the top 8 proteins was made using data from three individual studies conducted in Thailand, Brazil and the Solomon Islands (Longley et al., 2020); this was to identify markers that work well across multiple geographic regions, and not just in Thailand, for example.

For all 23 *P. vivax* proteins, we observed higher total IgG levels in individuals with current *P. vivax* infections compared to those without. This is again expected given these proteins were chosen due to their ability to identify individuals with recent past exposure to *P. vivax* parasites. We observed no relationship between *P. vivax* antigenic density and IgG level. A plausible reason for this finding, in contrast to our previous results (Longley et al., 2017b), is that all infections were asymptomatic and of relatively low parasite density. In addition, another key difference between the studies is that the present uses cross-sectional data (exact acquisition date of infection unknown) compared to an enrolled clinical cohort in our prior study (rough infection date known). What was perhaps unexpected was the statistically significant association of higher total IgG levels against all proteins in individuals with *P. falciparum* infections. This association remained even after the 11 individuals with co-infections were excluded. Potential reasons include cross-reactivity between orthologs in *P. vivax* and *P. falciparum*, or more simply because individuals with *P. falciparum* infections are also at risk of past *P. vivax* infections (as evidenced by recurrent *P. vivax* infections following treatment of *P. falciparum* infections (Douglas et al., 2011)). As only 9 of the 23 *P. vivax* proteins have *P. falciparum* orthologs it is the latter explanation that seems most likely. Increased risk of prior exposure to *Plasmodium* parasites is also likely the reason for the initially counter-intuitive finding of higher IgG levels in individuals using bednets. Previous analysis of the epidemiological data from this cross-sectional survey demonstrated that bednets had no protective effect in this population (Nguitragool et al., 2017), potentially due to the early outdoor biting phenotype of some *Plasmodium* vectors in Thailand (Sriwichai et al., 2016). It is plausible that the individuals who chose to use bednets did so because of past illness due to *Plasmodium* infections, thus they may be a group at higher risk of recent past exposure, potentially explaining the higher IgG levels. It is also likely that the malaria control program targeted distribution of bednets to where they were most needed, thus bednet usage could be considered a marker of where there is transmission.

A known risk factor for *Plasmodium* infections in Thailand is gender, with adult males being at higher risk of infection due to their occupation (Nguitragool et al., 2017). This increased risk of infection was also reflected in IgG levels to our 23 *P. vivax* proteins, which were generally higher in adults over 18 years of age and in men. The individuals in this study lived in three main regions, Bongti, Kong Mong Tha and Suan Phueng. *P. vivax* infections rates are highest in Bongti, and this was again reflected in our IgG measurements: IgG levels were highest in Bongti out of the three regions. Together, these analyses demonstrate that our panel of *P. vivax* serological exposure markers can be applied in a low-transmission setting to identify both individuals at high risk of infection and spatial clustering of infections at the region level.

We finally used a Random Forest algorithm that incorporated the antibody responses against the top 8 *P. vivax* serological exposure markers to predict which individuals had recent exposure to the parasite in the past 9 months. We used a cut-off prediction value to prioritise a high specificity target (i.e. 90% specificity, 62% sensitivity, Figure S1). A high specificity target for the serological exposure marker tool is better suited for surveillance to identify hot-spots or clusters of infections. As the number of true recent infections is low, it is important to have a high specificity rate to not over-estimate the levels of recent transmission. Once those areas have been identified, it would then be appropriate to use serological testing and treatment (seroTAT) as an intervention to identify hypnozoite carriers who are often asymptomatic. This will result in some misclassification, but as we previously demonstrated through modelling (Longley et al., 2020), is preferable to both mass drug administration (MDA) to avoid over-treatment and mass screening and treatment (using currently available diagnostics, i.e. microscopy or RDT) with poorer sensitivity. With the 62% sensitivity and 90% specificity target used in the current study, we identified a total 664 sero-positive individuals with an estimated 15.6% sero-prevalence. This included 108 out of the 144 individuals with PCR confirmed *P. vivax* infections. After adjusting the sero-prevalence estimate for the known sensitivity and specificity, we calculate an adjusted sero-prevalence of 10.8%. While the true cumulative exposure to infectious bites in the last 9 months is unknown in this population, we can infer from a longitudinal study in Thailand with similar PCR prevalence (mean 3.4%, range 1.7–4.2% (Nguitragool et al., 2019)), that the cumulative PCR prevalence over 9 months was 10%. Since our study was conducted earlier (2012 vs. 2013-2014) and during the high season, we expect slightly higher transmission levels and higher cumulative exposure, and therefore our adjusted estimate falls within the range that we would expect in this setting. We observed a significant moderate correlation between village-level PCR prevalence and the adjusted sero-prevalence from the algorithm, again demonstrating that our markers can be used for surveillance to determine clustering or hot-spots of infection.

Two systematic reviews on serological responses to *Plasmodium* proteins have highlighted that a) serological markers of surveillance could be used as an important and reliable tool for assessing malaria endemicity and exposure history, and b) that key knowledge gaps remain, particularly concerning the appropriate *P. vivax* proteins to be used and studies outside the South American region (Cutts et al., 2014;Folegatti et al., 2017). There have been a limited number of prior studies using detection of antibody responses against various *P. vivax* proteins as an indication of past exposure in the Asia-Pacific region; these include two studies conducted in Cambodia and Vanuatu. These studies have mainly used responses against relatively well-known proteins, such as the circumsporozoite protein (CSP), MSP1 (which is also in our 8-protein panel), the Duffy binding protein (DBP, in our larger panel) and apical membrane antigen 1 (AMA1) (Kerkhof et al., 2016;Idris et al., 2017). A challenge highlighted was the relatively long estimated antibody half-life against most *P. vivax* proteins tested (Kerkhof et al., 2016), suggesting it would be difficult to distinguish between recent and long-term changes in transmission patterns. Indeed the study in Vanuatu was aiming to demonstrate changes in transmission over a 4-year period (Idris et al., 2017), which is useful for monitoring endemicity and demonstrating the power or utility of various interventions. However, long-term changes in transmission are not useful for applying sero-surveillance as an intervention in itself. The power of our specific panel of proteins is that they were carefully selected to reflect recent exposure to *P. vivax* parasites, rather than lifetime exposure, in low-transmission settings. Furthermore, using the combination of IgG responses to our top 8 proteins enables classification of recent infections with high accuracy and also introduces flexibility to account for potential differences in antibody half-life and seropositivity rates in different geographic regions. We have demonstrated the power of our markers for surveillance and identifying hot-spots of infections. These hot-spots can then be further targeted using our markers in a seroTAT approach, where all sero-positive individuals could be treated with anti-malarial drugs. Compared to PCR, we identified additional individuals likely harbouring hypnozoites who need to be targeted for *P. vivax* transmission to be interrupted.

## Data Availability

All datasets for this study are included in the manuscript and the supplementary files.

## 5 Supplementary Material

Table S1: Complete data set (antibody responses and epidemiological variables).

Table S2: Univariable and multivariable analysis of antibody responses with epidemiological or demographic factors.

Figure S1: Random Forest classification algorithm receiver operator characteristic curve (ROC).

## 7 Author Contributions

IM, JS and RL designed the study. CK, KK, PS, WN and JS conducted the cross-sectional survey. ET, TT, W-HT, MH, CC and JH expressed proteins. JB and RL performed protein coupling. SC, CK, JH performed antibody measurements. SC and RL did the data management and analysis. MW and NN performed statistical modeling. SC and RL wrote the first draft of the manuscript. All authors contributed to data interpretation and revision of the manuscript.

## 9 Acknowledgments

We gratefully acknowledge the extensive field teams that contributed to sample collection and qPCR assays. We thank all individuals that participated in the cross-sectional survey. We thank Connie Li-Wai-Suen for writing the R script used for converting MFI values to relative antibody units. We thank Christopher King (Case Western Reserve University) for provision of the PNG control plasma pool.

## 11 Data availability statement

All datasets for this study are included in the manuscript and the supplementary files.

