## Supplementary figures and images for "Application of 23 novel serological markers for identifying recent exposure to *Plasmodium vivax* parasites in an endemic population of western Thailand"

### Supplemental Figure 1

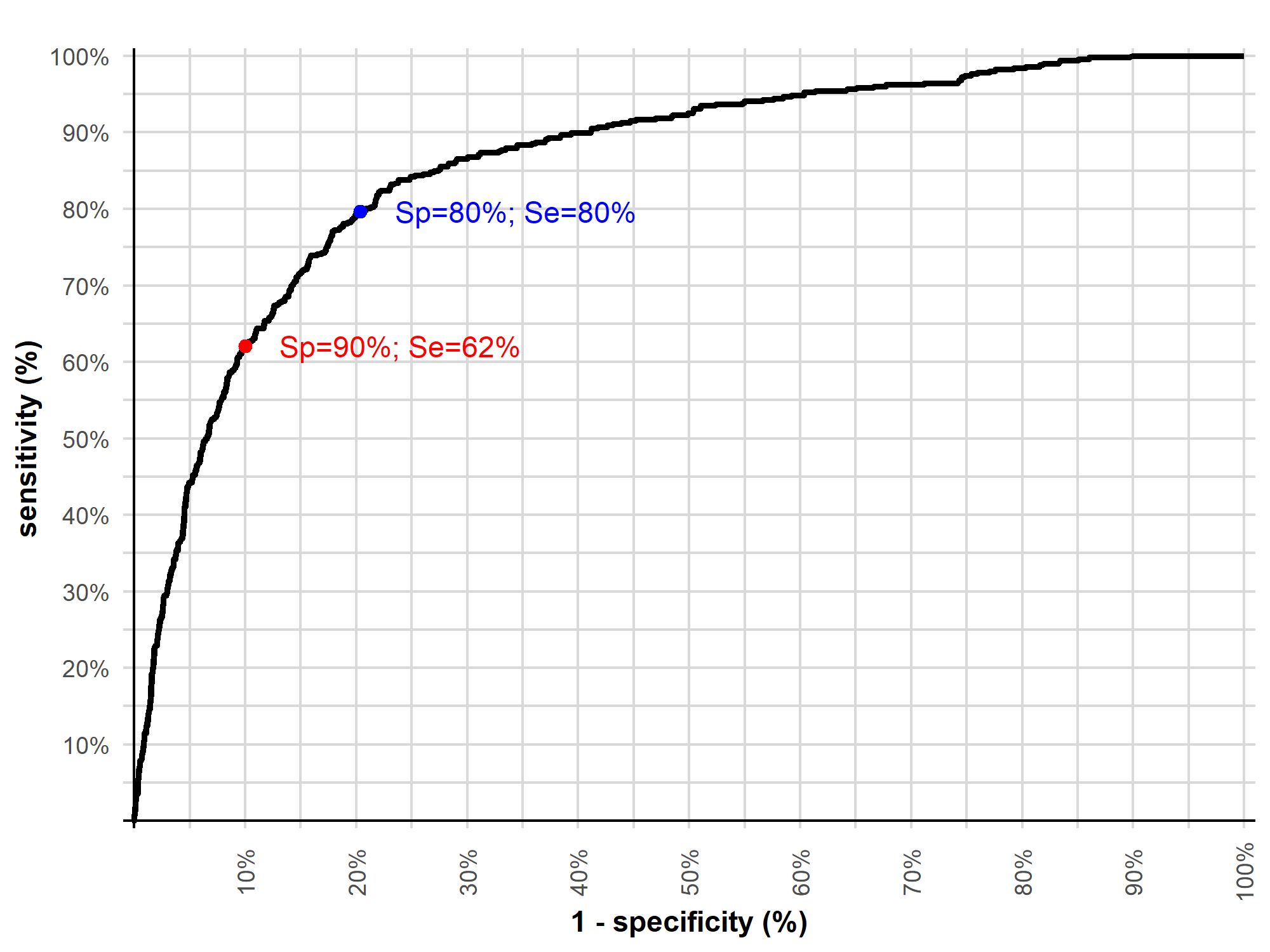
